# Developing and Evaluating Deep Learning Approaches for Visual Field Denoising in Glaucoma

**DOI:** 10.64898/2026.05.29.26354019

**Authors:** Julia Seungjoo Baek, Anagha Lokhande, Didier Neuenschwander, Min Shi, Mengyu Wang

## Abstract

**Purpose:** To investigate the relative efficacy of nine distinct visual field (VF) denoising artificial intelligence (AI) methods and a pathology-aware AI strategy to discourage over-correction of glaucomatous defects.

**Design:** Retrospective study

**Participants:** 87,940 paired visual field (VF) and optical coherence tomography (OCT) samples from a tertiary academic center.

**Methods:** Denoising models were trained on a separate VF-only dataset and evaluated on an independent structure-function dataset of paired VF-OCT samples. We implemented and evaluated nine distinct VF denoising strategies representing three broad categories: baseline measurements, self-supervised and image restoration models (including Noise2Noise, Noise2Void, and NAFNet), and latent variable compression-based models (autoencoders and variational autoencoders). All models were designed to reconstruct VF sensitivity maps. We then predicted retinal nerve fiber layer thickness (RNFLT) maps from the denoised VFs using a fixed, independently trained VF-to-RNFLT prediction model.

**Main Outcome Measures:** Predicted VF and RNFLT maps and resultant evaluation metrics.

**Results:** The raw VF baseline achieved a global R² of 0.5468 and MAE of 16.83 μm. Restoration-based models maintained or slightly improved concordance, with the pathology-aware NAFNet achieving the highest global R² (0.5485) and a comparable MAE (16.82 μm). In contrast, compression-based models degraded concordance, with CNN-VAE showing a significant reduction (R² ≈ 0.50).

In severe glaucoma, concordance decreased across all methods; however, compression architectures exhibited disproportionately greater degradation compared with restoration-based approaches.

**Conclusions:** We present a comparative benchmark of AI-based VF denoising strategies paired with structure–function evaluation. While restoration-based models can reduce variability without loss of biological signal, latent compression risks attenuating clinically meaningful defects. Visually smoother fields are not necessarily more biologically accurate.

## Introduction

Glaucoma is a leading cause of irreversible blindness worldwide and is characterized by progressive loss of retinal ganglion cells (RGCs) and corresponding visual field (VF) defects.^1^ Standard Automated Perimetry (SAP) remains the primary clinical tool for assessing functional impairment in glaucoma.^2^ However, its utility is limited by substantial measurement variability arising from patient fatigue, fixation instability, and the psychophysical nature of the test. This variability complicates the detection of disease progression and weakens the relationship between functional loss and underlying structural damage.^3^

Recent studies have applied deep learning methods to denoise visual field data.^4–7^ Techniques ranging from variational autoencoders to convolutional neural networks have demonstrated the ability to smooth VF maps and reduce apparent noise. Despite these advances, a critical validation challenge persists. In clinical practice, the true noise-free visual field is unknown. Without a ground truth reference, it is difficult to determine whether denoising enhances signal fidelity or inadvertently obscures subtle pathological defects.

To address this limitation, we propose a validation framework grounded in the glaucoma structure–function relationship.^8,9^ Because functional loss in glaucoma arises from retinal ganglion cell degeneration and retinal nerve fiber layer (RNFL) thinning, improved denoising should yield stronger agreement between functional and structural measurements. Structure–function mapping therefore provides a biologically meaningful proxy for evaluating denoising performance.

In this study, we benchmark nine distinct visual field denoising strategies using structure–function agreement as an objective standard. Furthermore, we introduce a pathology-aware training strategy applied to a NAFNet architecture that incorporates severity-dependent and spatially informed loss weighting to discourage over-smoothing of deep glaucomatous defects. This approach aims to preserve clinically meaningful scotomas while reducing measurement noise.

## Methods

### Data Collection

A VF-only dataset from the Glaucoma Research Network (GRN) consortium was used exclusively to train the denoising models. This dataset was kept strictly separate from the structure–function evaluation dataset to prevent data leakage. The raw dataset contained 471,527 VF tests from 204,619 eyes of 116,288 patients. This study was approved by the institutional review boards of all participating institutions and adhered to the tenets of the Declaration of Helsinki and the United States Health Insurance Portability and Accountability Act of 1996.

Duplicate entries were identified and removed based on patient ID, eye, age, test duration, and total deviation values, reducing the dataset to 452,317 records. A standard reliability filter was then applied, retaining only examinations with fixation losses ≤ 33%, false negatives ≤ 20%, and false positives ≤ 20%. This resulted in a final filtered training dataset of 394,083 tests from 180,524 eyes of 103,136 distinct patients. The mean patient age in this filtered dataset was 62.2 ± 16.6 years, with a mean of 2.18 exams per eye.

Structure–function concordance was evaluated using a refined subset of the Massachusetts Eye and Ear Glaucoma Service testing dataset. Initially, the raw clinical dataset comprised 416,367 Humphrey 24-2 visual field (VF) examinations. After applying reliability filters (fixation losses ≤ 33%, false positive and false negative rates ≤ 20%), the dataset consisted of 293,348 records representing 73,152 distinct patients and 142,431 distinct eyes.

For the final evaluation set, these records were further restricted to those with paired Cirrus OCT RNFL thickness maps (signal strength ≥ 6) obtained within a ±3-month window (|Δt| ≤ 90 days) of the corresponding VF. Following the exclusion of 5,484 samples exhibiting RNFL thickness artifacts exceeding 350 μm, the final paired dataset included 87,940 VF–OCT pairs from 41,433 eyes of 24,422 patients. Patients were partitioned at the patient level into training (80%), validation (10%), and test (10%) sets to prevent data leakage. The held-out test set consisted of 8,794 samples from 7,819 eyes of 6,875 patients and was used for all final performance metrics.

To satisfy the specific architectural requirements of the different denoising models, the training data were partitioned based on the need for temporal test-retest series. The datasets were assigned as follows:

- **Noise2Void (N2V):** As a blind denoising method requiring only single independent visual fields, N2V did not utilize test-retest grouping and was trained on the entire filtered dataset of 394,083 individual tests (N ≥ 1).
- **Noise2Noise (N2N):** Because N2N requires paired noisy measurements to suppress uncorrelated noise effectively, it was trained on a specific subset of eyes with at least two reliable tests (N ≥ 2) acquired within a 90-day span, comprising 28,863 samples.
- **NAFNet and Latent Compression Models:** The pathology-aware NAFNet and all compression-based architectures (CNN-VAE, NN-AE, Vanilla AE, VAE, PosEnc) were trained using a more rigorous subset of eyes with at least three reliable tests (N ≥ 3) within a 90-day span, comprising 2,769 samples. For the compression-based models, this specific subset provided the necessary proxy targets to support their two-stage training procedure.

### Denoising Models

We implemented and evaluated nine distinct visual field denoising strategies representing three broad categories: baseline measurements, self-supervised and image restoration models, and latent variable compression-based models. All models were designed to reconstruct visual field sensitivity maps and functioned as content-aware filters intended to suppress measurement noise while preserving glaucomatous patterns.

**Table 1.** Overview of visual field denoising baseline models. Abbreviations: TD = total deviation; VF = visual field; VAE = variational autoencoder; AE = autoencoder; MSE = mean squared error; KL = Kullback–Leibler; N2N = Noise2Noise; N2V = Noise2Void.

| Model | Category | Input Representation | Key Characteristics |
| --- | --- | --- | --- |
| Raw Visual Field | Baseline | 52 TD values | Unprocessed clinical visual field measurements used as the reference condition without denoising. |
| Noise2Noise (N2N) <sup>5</sup> | Self-supervised denoising | 2D VF grid | Trained on paired noisy VF measurements from repeated examinations. Learns the consistent underlying signal shared across visits while suppressing random perimetric variability. |
| Noise2Void (N2V) <sup>6</sup> | Blind denoising | 2D VF grid | Masked prediction of individual test locations using surrounding spatial context. Removes isolated noise assuming independence between measurement errors. |
| NAFNet <sup>4</sup> | Image restoration | 2D VF grid | Activation-free architecture optimized for high-fidelity image restoration. Designed to preserve sharp spatial gradients corresponding to nerve fiber bundle defect borders. |
| Pathology-Aware NAFNet | Image restoration | 2D VF grid | Modified reconstruction loss emphasizing deep scotomas and clinically relevant VF regions to prevent attenuation of advanced glaucomatous defects. |
| MLP Autoencoder | Compression model | 52-dimensional TD vector | Fully connected encoder-decoder architecture compressing VF data into latent space ($z=8$) with MSE reconstruction loss. |
| CNN Autoencoder | Compression model | 12×12 VF grid + mask | Convolutional encoder-decoder using masked MSE to reconstruct only valid test locations. |
| MLP Variational Autoencoder | Compression model | 52-dimensional TD vector | Probabilistic latent representation with KL divergence regularization combined with reconstruction loss. |
| CNN Variational Autoencoder | Compression model | 12×12 VF grid + mask | Convolutional VAE capturing spatial VF structure while regularizing latent distribution. |
| Positional Attention Autoencoder | Compression model | 52 TD values + positional encoding | Incorporates sinusoidal positional embeddings and self-attention to model non-local spatial dependencies before latent compression. |

For denoising models requiring paired noisy and clean targets, specifically the pathology-aware NAFNet and all latent compression-based architectures, a test-retest subset was used in which each eye had at least three reliable HFA 24-2 examinations acquired within a 90-day span. After confirming spatial alignment of test locations across visits, an arithmetic per-location average of the total deviation (TD) values was computed and treated as a denoised proxy target. All examinations used to generate this averaged target were acquired within a 90-day interval to minimize the influence of true disease progression. A single-visit visual field from this series served as the corresponding noisy input. This strategy exploits the incoherent nature of perimetric noise, as averaging repeated measurements suppresses random fluctuations while preserving consistent pathological signal. Self-supervised and blind denoising models, including Noise2Noise and Noise2Void, were trained without explicit clean targets using their respective training paradigms.

The original, unprocessed total deviation (TD) values from standard automated perimetry served as the baseline reference. These values represent deviation from age-matched normative sensitivity at each test location and reflect the inherent structure–function relationship present in routine clinical testing without mathematical denoising.

Supervised image restoration and self-supervised models treat the visual field as a two-dimensional image and attempt to separate biological signal from measurement noise arising from random perimetric fluctuation.^10,11^ Models were trained on the dedicated VF-only training dataset and subsequently applied to infer denoised sensitivity maps in the independent structure–function evaluation dataset.^12,13^

Compression-based models assume that glaucomatous visual field patterns lie on a lower-dimensional manifold relative to random perimetric noise. Visual fields were compressed into a latent representation prior to reconstruction. All TD values were normalized to [-1, 1], and reconstructed outputs used a final hyperbolic tangent activation to preserve scaling.^11,14^ All compression-based models were trained using a two-stage training strategy. In the first stage, the full encoder–decoder network was optimized end-to-end using reconstruction loss. In the second stage, early encoder layers were frozen while the decoder and final projection layers were fine-tuned to improve reconstruction fidelity while preserving the learned latent manifold.

### Pathology-Aware Training Strategy

To reduce the risk of pathological signal attenuation during denoising, we implemented a pathology-aware training strategy incorporating both severity-dependent and spatially informed loss weighting.

First, reconstruction errors were weighted according to the degree of functional depression at each visual field location. Test points with greater sensitivity loss contributed proportionally more to the overall loss, thereby emphasizing preservation of deep scotomas and steep spatial gradients characteristic of glaucomatous damage. This severity-aware weighting was designed to counteract the tendency of neural networks to regress extreme values toward the population mean (Supplemental Figure 1).

Second, a static spatial weighting matrix was applied to prioritize anatomically and clinically informative regions of the visual field, including central and arcuate nerve fiber bundle zones. This matrix reflected the known topographic organization of retinal ganglion cell axon trajectories, utilizing principles such as the Garway-Heath mapping,^8^ and aimed to preserve spatially meaningful defect patterns. Finally, the training dataset was balanced through oversampling of moderate-to-severe glaucoma cases. This ensured adequate representation of advanced pathological patterns during optimization and reduced bias toward normal or mildly affected fields. Collectively, these modifications were designed to discourage over-smoothing and attenuation of true glaucomatous defects while maintaining suppression of stochastic perimetric noise.

To evaluate denoised visual fields using a biologically grounded criterion, structure–function agreement was quantified by predicting peripapillary RNFLT maps from visual field inputs.^9^ A separate VF-to-RNFLT model was trained independently of all denoising models and subsequently fixed as an evaluation framework. Each HFA 24-2 visual field was represented as a 52-point total deviation (TD) vector. To preserve spatial relationships while maintaining the original sampling geometry, the 52 test locations were mapped onto an 8×9 spatial grid corresponding to the 24-2 layout, with invalid corner locations left empty. During training of the VF-to-RNFLT model, the encoder component of the RNFLT autoencoder served as a fixed teacher network to generate normalized targets from ground-truth RNFLT maps. The pretrained RNFLT decoder was reused to reconstruct full-resolution RNFLT maps from predicted latent vectors. The VF-to-RNFLT model consisted of a visual field spatial encoder followed by the pretrained RNFLT decoder. The VF encoder comprised two convolutional layers operating on the 8×9 grid, followed by fully connected layers that projected the representation into the normalized RNFLT latent space. The predicted latent vector was rescaled using the training-set latent mean and standard deviation and then passed through the RNFLT decoder to generate a reconstructed RNFL thickness map. Latent-space error was computed as a mean squared error (MSE) between predicted and teacher latent vectors. The total loss was defined as a weighted combination of latent and pixel reconstruction terms. To preserve the learned RNFLT manifold and stabilize optimization, pretrained RNFLT encoder parameters were frozen. Only the VF encoder and final decoder layers were fine-tuned. Optimization was performed using the Adam optimizer with a validation-based learning rate scheduler.

**Figure 1.**
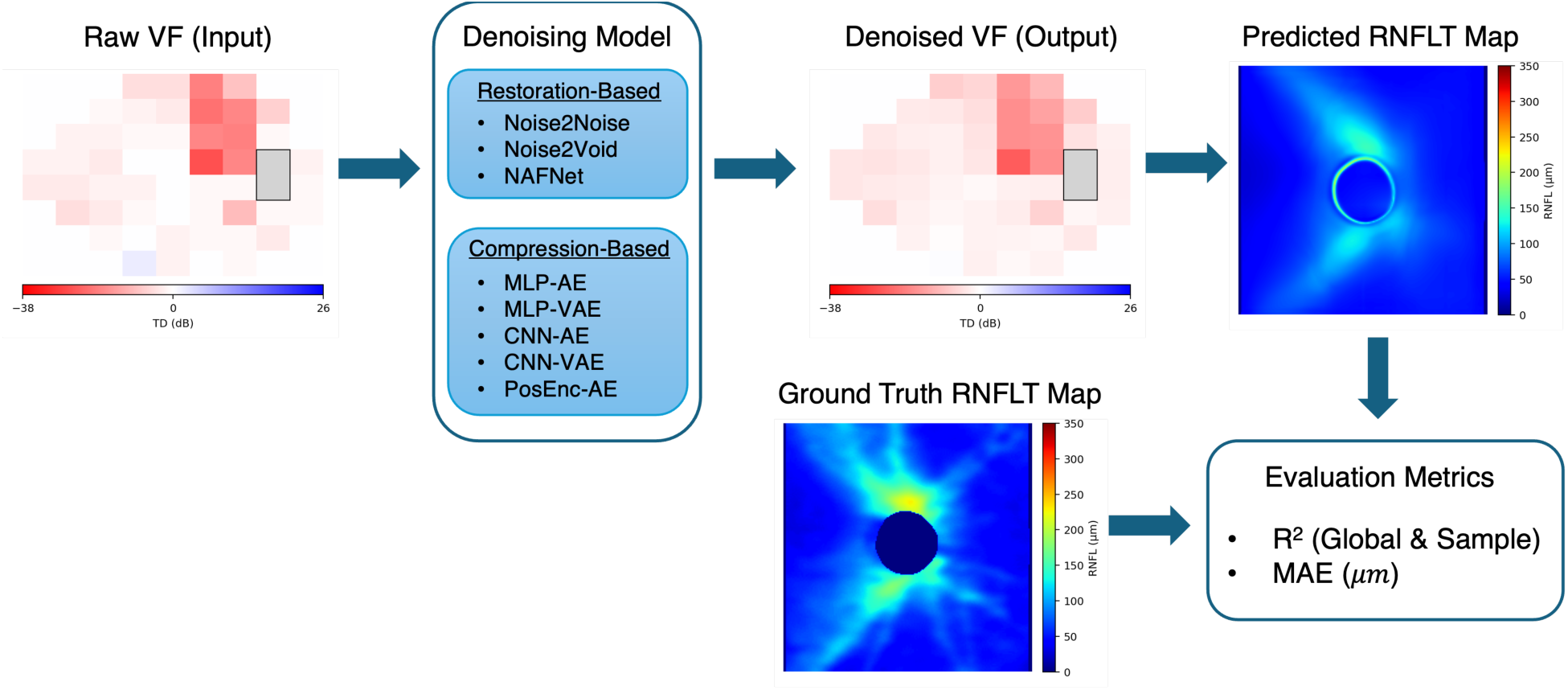
Overview of the evaluation framework. Raw visual fields (VF) are processed using restoration-based and compression-based denoising models. The denoised VF is then input to a fixed VF-to-RNFLT model to generate predicted RNFLT maps, which are compared against ground-truth OCT RNFLT maps using MAE (μm) and R² within the circular masked region.

### Evaluation Metrics

For each denoising strategy, denoised visual fields were provided as input to the fixed VF-to-RNFLT model. Predicted RNFLT maps were compared with corresponding OCT-derived RNFL thickness maps on a held-out test set. Performance was quantified using mean absolute error (MAE, 𝜇m) and coefficient of determination (R^2^) computed within the circular masked region.

Higher R^2^ and lower MAE were interpreted as stronger structure–function concordance, indicating improved preservation of functional information relevant to retinal nerve fiber layer structure.

### Statistical Analysis

Performance was evaluated globally and stratified by disease severity based on Mean Deviation (MD):

- **Borderline/Normal:** MD ≥−3 dB
- **Mild:** −6≤ MD <−3 dB
- **Moderate:** −12≤ MD <−6 dB
- **Severe:** MD <−12 dB

Global metrics were computed across all test samples. Subgroup analyses were performed within each severity category to assess whether denoising effects differed by stage of disease. All statistical evaluations were conducted on a held-out test set not used during training of either denoising models or the VF-to-RNFLT predictor. The VF-to-RNFLT model remained fixed across all comparisons to ensure methodological consistency.

To assess whether evaluation direction influences sensitivity to spatial information preservation, two additional analyses were performed. First, an RNFLT-to-VF prediction model was trained and evaluated using each denoising strategy as input. An averaged-VF condition was also constructed in which all 52 test locations were replaced by the mean sensitivity value, thereby preserving global severity while eliminating spatial structure. Second, the VF-to-RNFLT evaluation pipeline was repeated using averaged visual fields as input. This allowed direct assessment of whether spatial collapse degraded structure–function agreement in the primary evaluation direction. These experiments were designed to determine whether each evaluation framework was sensitive to preservation of spatially localized glaucomatous defects.

## Results

The held-out test cohort consisted of 8,794 visual field examinations from 7,819 eyes of 6,875 patients. The mean patient age at the time of testing was 61.2 ± 15.3 years (range, 10.1–99.1), and 57.7% of patients were female. The racial distribution of the cohort was 66.2% White or Caucasian (n=4,549), 13.5% Black or African American (n=926), and 8.7% Asian (n=595). Smaller proportions of patients were classified as Other (7.0%, n=479), Unknown or Missing (4.3%, n=298), American Indian or Alaska Native (0.2%, n=15), Hispanic or Latino (0.1%, n=7), and Native Hawaiian or Other Pacific Islander (0.1%, n=6). Regarding ethnicity, 8.4% of patients were identified as Hispanic (n=579), while 87.6% were non-Hispanic (n=6,024); ethnicity information was missing for 4.0% of patients (n=272). Based on mean deviation (MD), glaucoma severity among eyes in the test set was distributed as follows: borderline or normal in 5,086 eyes (65.0%), mild in 1,464 eyes (18.7%), moderate in 794 eyes (10.2%), and severe in 475 eyes (6.1%). A summary of cohort characteristics is provided in Table 2.

**Table 2.** Characteristics of the held-out test cohort.

| Characteristic | Value |
| --- | --- |
| Number of scans | 8,794 |
| Number of eyes | 7,819 |
| Number of patients | 6,875 |
| Age, mean ± SD (range), years | 61.2 ± 15.3 (10.1–99.1) |
| Sex, n (%) |  |
| Male | 2,910 (42.3%) |
| Female | 3,965 (57.7%) |
| Race, n (%) |  |
| White or Caucasian | 4,549 (66.2%) |
| Black or African American | 926 (13.5%) |
| Asian | 595 (8.7%) |
| Other | 479 (7.0%) |
| Unknown / Missing | 298 (4.3%) |
| American Indian or Alaska Native | 15 (0.2%) |
| Hispanic or Latino | 7 (0.1%) |
| Native Hawaiian or Other Pacific Islander | 6 (0.1%) |
| Hispanic ethnicity, n (%) |  |
| Hispanic | 579 (8.4%) |
| Non-Hispanic | 6,024 (87.6%) |
| Missing | 272 (4.0%) |
| Glaucoma severity (per eye), n (%) |  |
| Borderline | 5,086 (65.0%) |
| Mild | 1,464 (18.7%) |
| Moderate | 794 (10.2%) |
| Severe | 475 (6.1%) |

### Global Structure–Function Agreement

VF-to-RNFLT predictive performance was evaluated across all nine denoising strategies on the held-out test set (n=8,794; Table 3). Representative denoising outputs are shown in Figure 2, highlighting qualitative differences in defect morphology preservation across restoration-based and compression-based approaches. To assess whether these qualitative differences translated into structural correspondence, we compared RNFLT maps predicted from denoised visual fields against OCT-derived RNFLT maps (Figure 3). The raw visual field baseline achieved a global R^2^ of 0.5468 with a mean absolute error (MAE) of 16.83 ± 5.43 μm (Table 3).

**Figure 2.**
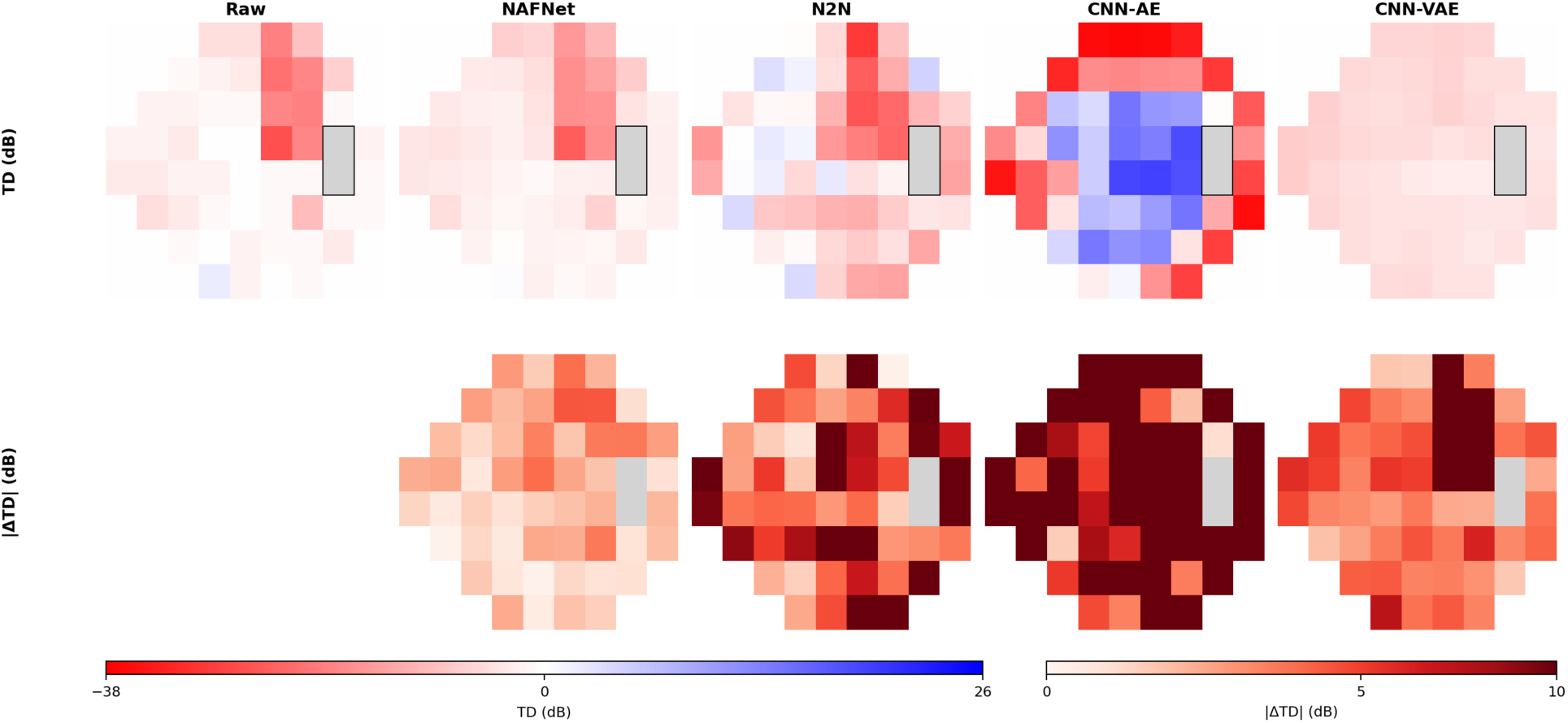
Representative visual field denoising results. Visual fields from the same eye were denoised using representative restoration-based and compression-based models. Compression-based architectures attenuated deep scotomas and reduced spatial contrast, whereas restoration-based approaches preserved localized defect morphology. All panels use an identical color scale.

**Figure 3.**
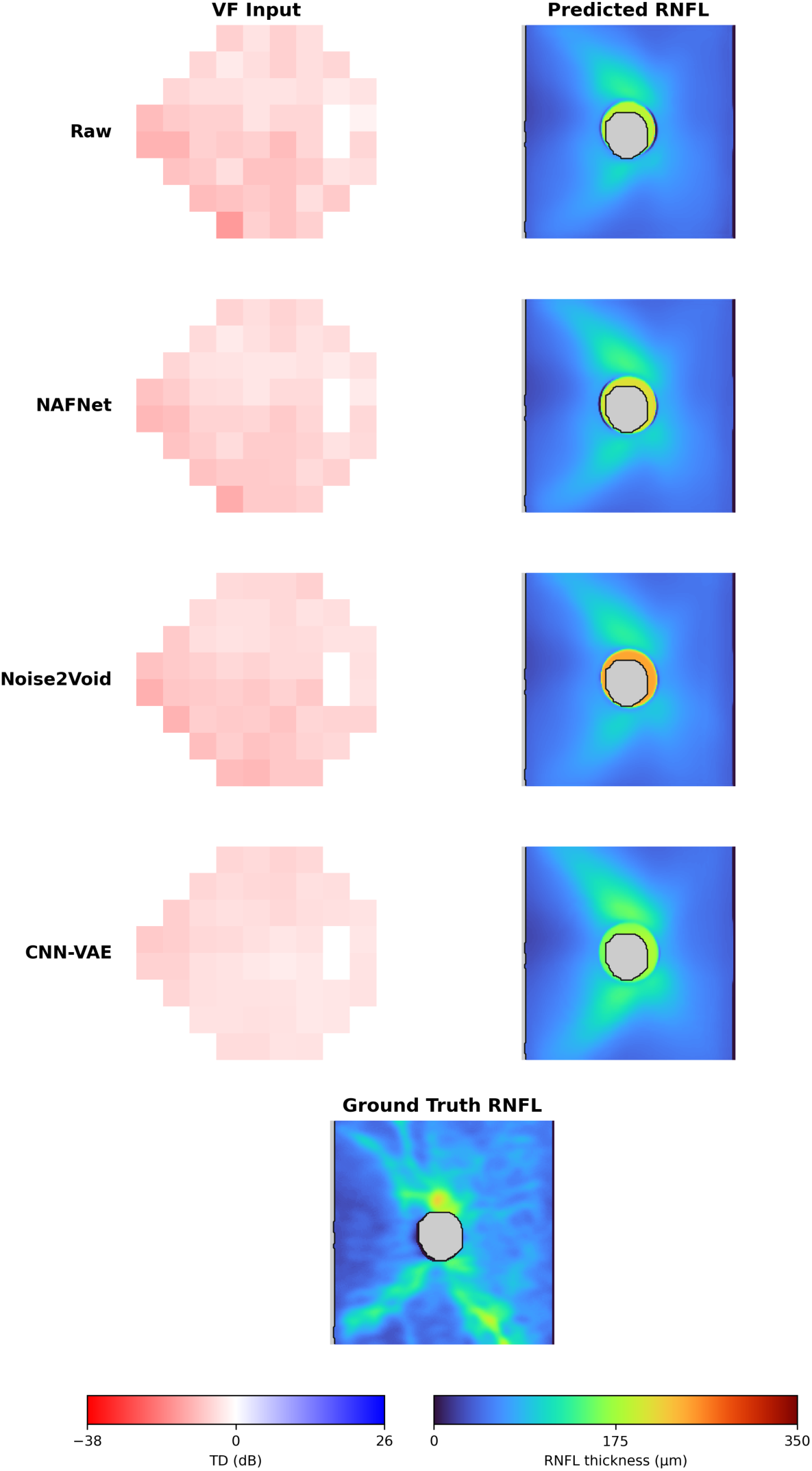
Effect of denoising on structure–function mapping. Predicted RNFLT maps generated from denoised visual fields were compared with OCT-derived RNFLT maps. Although global RNFLT patterns were broadly preserved across models, compression-based denoising resulted in reduced focal structural contrast, whereas pathology-aware NAFNet maintained closer alignment with localized structural damage.

**Table 3.** Global VF-to-RNFLT performance on the held-out test set (n=8,794).

| Model | Global R<sup>2</sup> | MAE (μm) |
| --- | --- | --- |
| Raw Visual Field | 0.5468 | 16.83 ± 5.43 |
| Restoration-based |  |  |
| NAFNet (Pathology-Aware) | 0.5485 <sup>a</sup> | 16.82 ± 5.45 <sup>a</sup> |
| Noise2Void (N2V) | 0.5438 | 16.87 ± 5.57 |
| Noise2Noise (N2N) | 0.5283 | 17.26 ± 5.57 |
| Compression-based |  |  |
| CNN-AE | 0.5388 | 16.94 ± 5.58 |
| Vanilla AE (MLP) | 0.5387 | 16.97 ± 5.57 |
| VAE (MLP) | 0.5208 | 17.34 ± 5.64 |
| CNN-VAE | 0.5045 | 17.69 ± 5.58 |
| PosEnc-AE | 0.5423 | 16.91 ± 5.53 |

<sup>a</sup> denotes best performance within category.

Among restoration-based approaches, pathology-aware NAFNet achieved the highest global R^2^ (0.5485) with an MAE of 16.82 ± 5.45 μm (Table 3). Consistent with the preserved defect morphology observed in Fig. 2, NAFNet maintained localized structure–function alignment in predicted RNFLT maps (Figure 3). Noise2Void (N2V) and the Positional Encoding Autoencoder (PosEnc) achieved global R^2^ values of 0.5438 and 0.5423, respectively, whereas Noise2Noise (N2N) demonstrated reduced concordance (R^2^ = 0.5283).

In contrast, compression-based models demonstrated progressively lower structure–function agreement (Table 3). CNN-AE and Vanilla AE achieved global R^2^ values of 0.5388 and 0.5387, respectively. Variational architectures exhibited further degradation (VAE: 0.5208; CNN-VAE: 0.5045), with CNN-VAE demonstrating the largest reduction in concordance. Qualitatively, these models attenuated deep scotomas and reduced spatial contrast (Figure 2), which corresponded to diminished localized structural correspondence in predicted RNFLT maps (Figure 3).

### Performance by Disease Severity

Structure–function concordance declined progressively with increasing disease severity across all models. In borderline eyes, restoration-based approaches performed comparably to or slightly better than raw input (NAFNet: R^2^ = 0.5855 vs. raw: 0.5843). Similar trends were observed in mild and moderate subgroups, where pathology-aware NAFNet consistently achieved the highest concordance.

**Table 4.** R² stratified by disease severity (VF-to-RNFLT mapping).

| Model | Borderline<br>(n=5,752) | Mild<br>(n=1,646) | Moderate<br>(n=873) | Severe<br>(n=523) |
| --- | --- | --- | --- | --- |
| Raw | 0.5843 | 0.4885 | 0.4235 | 0.3357<sup>a</sup> |
| NAFNet (Pathology-Aware) | 0.5855<sup>a</sup> | 0.4942<sup>a</sup> | 0.4283<sup>a</sup> | 0.3212 |
| Noise2Void (N2V) | 0.5831 | 0.4854 | 0.4166 | 0.3084 |
| CNN-VAE | 0.5640 | 0.4529 | 0.3154 | 0.0092 |

<sup>a</sup> indicates highest value within each severity column.

In severe glaucoma, concordance declined across all models (raw: R^2^ = 0.3357), consistent with the known RNFLT floor effect.^15,16^ Restoration-based models did not improve performance beyond baseline in this subgroup. Compression-based architectures, particularly CNN-VAE (R^2^ = 0.0092), demonstrated marked degradation.

**Figure 4.**
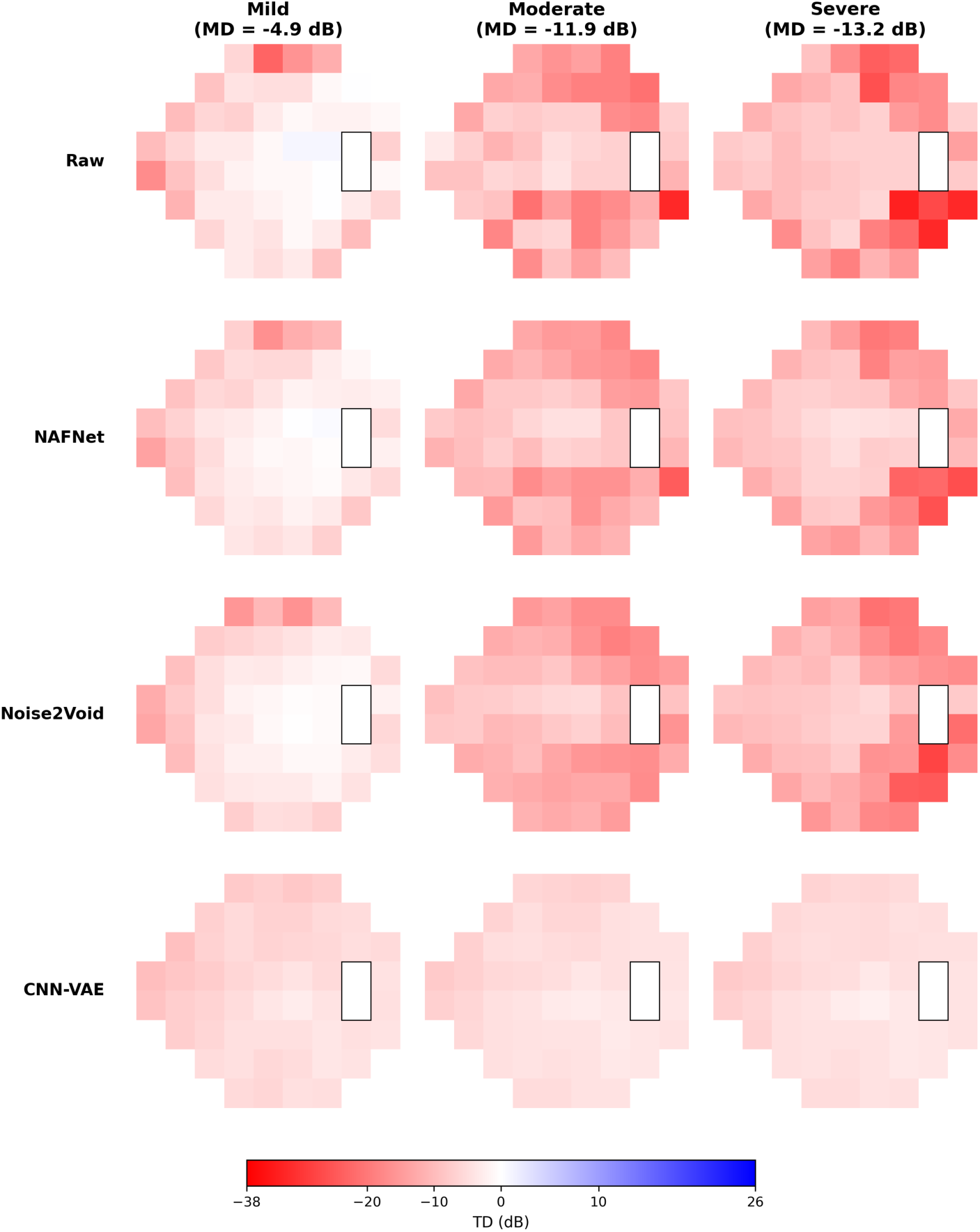
Severity-stratified visual field denoising examples. Representative mild, moderate, and severe glaucoma cases (Mean Deviation shown in column titles) comparing raw visual fields to restoration-based denoising (NAFNet, Noise2Void) and compression-based denoising (CNN-VAE). In severe glaucoma, CNN-VAE markedly attenuates deep scotomas and reduces spatial contrast, whereas restoration-based models better preserve localized defect morphology. All panels share an identical color scale.

### Evaluation Direction Sensitivity

To assess whether evaluation direction is sensitive to preservation of spatially localized defect morphology, we compared RNFLT-to-VF prediction with VF-to-RNFLT mapping under two input conditions: (i) spatially preserved visual fields and (ii) spatially collapsed inputs in which all 52 test locations were replaced by the mean sensitivity value (Averaged-VF). Because the Averaged-VF target is constant across locations, R^2^ and Pearson correlation are not defined for this condition; therefore, comparison relied on absolute error metrics (MAE and RMSE).

In the RNFLT-to-VF direction, predictive performance remained high across multiple denoising strategies even when spatial structure was removed. Notably, CNN-based compression models achieved near-ceiling performance with spatially preserved inputs (CNN-AE: R^2^=0.9735, Pearson=0.9932; CNN-VAE: R^2^=0.9381, Pearson=0.9835), consistent with strong reliance on global severity cues. In this direction, spatial averaging did not substantially degrade error and in several cases reduced it (Table 5), indicating relative insensitivity to spatial morphology.

**Table 5.** Evaluation-direction sensitivity in RNFLT-to-VF prediction. Spatially averaged visual fields remove defect morphology while preserving global severity. Mean absolute error (MAE) and root mean squared error (RMSE) are reported.

| Model | MAE<br>(Preserved) | MAE<br>(Averaged) | RMSE<br>(Preserved) | RMSE<br>(Averaged) |
| --- | --- | --- | --- | --- |
| Raw | 2.7813 | 2.0090 | 3.6308 | 2.0168 |
| PosEnc-AE | 2.0763 | 1.7482 | 2.4262 | 1.7578 |
| NAFNet | 2.2532 | 1.7806 | 2.8369 | 1.7903 |
| Noise2Void (N2V) | 2.3384 | 1.9639 | 2.8503 | 1.9776 |
| Noise2Noise (N2N) | 4.8215 | 2.4774 | 6.0867 | 2.4892 |
| MLP-AE | 2.0478 | 1.7820 | 2.3907 | 1.7975 |
| MLP-VAE | 1.7444 | 1.6565 | 1.8349 | 1.6665 |
| CNN-AE | 1.6375 | 1.3879 | 2.1787 | 1.3950 |
| CNN-VAE | 0.2200 | 0.1459 | 0.2727 | 0.1478 |

By contrast, the VF-to-RNFLT evaluation framework penalized spatial collapse: replacing each VF with its mean value reduced structure–function concordance (lower R^2^ and higher MAE), demonstrating substantially greater sensitivity to preservation of localized defect patterns.

Overall, RNFLT-to-VF prediction remained highly accurate in several models despite complete removal of spatial defect morphology, whereas VF-to-RNFLT mapping demonstrated clear sensitivity to spatial collapse. These findings indicate that evaluation direction fundamentally influences apparent denoising performance, afnd that anatomically constrained VF-to-RNFLT mapping provides a more spatially discriminative validation framework.

**Figure 5.**
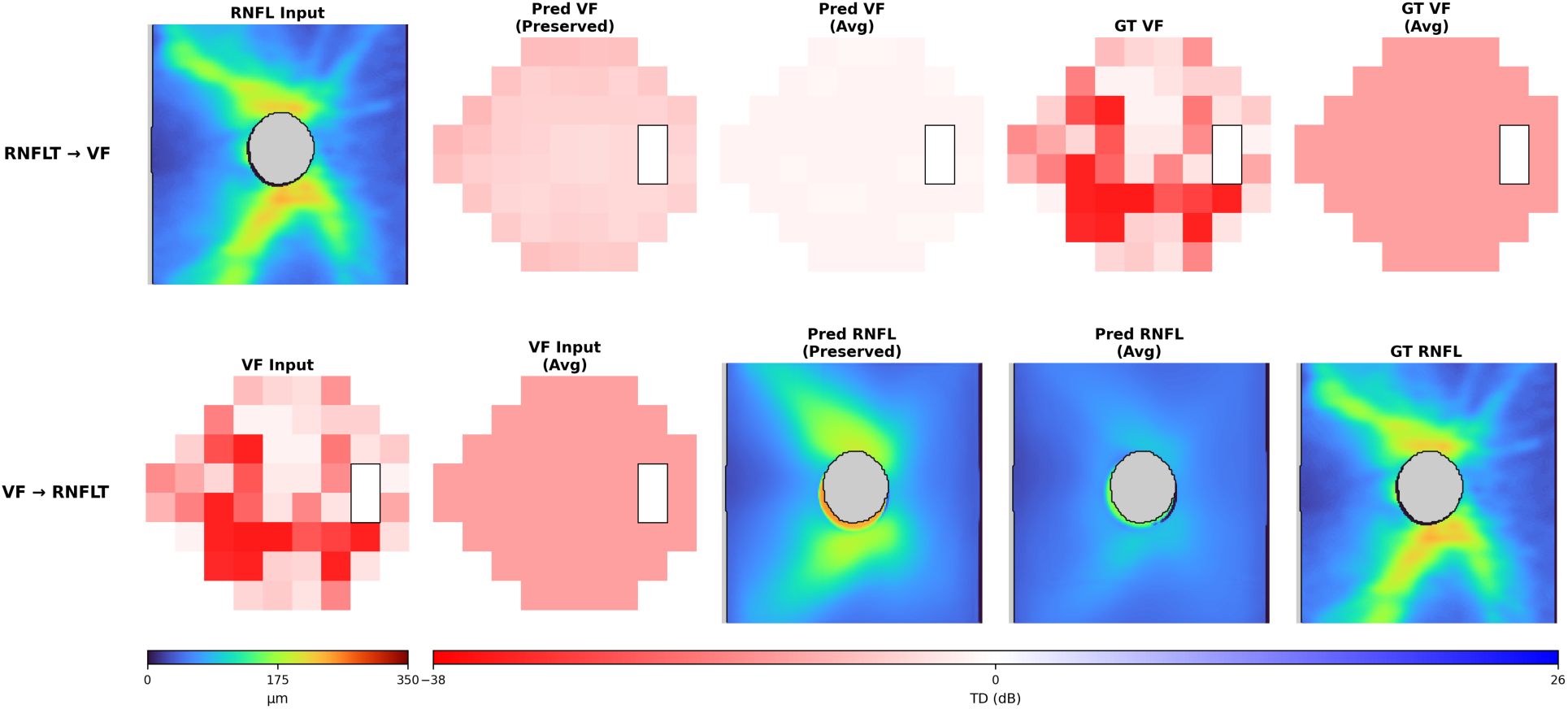
Directional sensitivity to spatial collapse. (A) In the RNFLT→VF direction, spatial averaging of VF inputs removes localized defect morphology but preserves global severity, allowing models to maintain low error despite loss of spatial structure. (B) In the VF→RNFLT direction, spatial averaging eliminates critical localized scotomas required for structural localization, resulting in degraded RNFLT prediction. These examples illustrate why RNFLT-to-VF evaluation may overestimate denoising fidelity when models rely predominantly on global severity cues.

## Discussion

To our knowledge, this study represents the largest comparative evaluation of deep learning–based visual field denoising strategies assessed using structure–function mapping as a biologically grounded validation framework. Our principal finding is that most denoising approaches do not meaningfully improve structure–function concordance beyond the raw visual field baseline, and that certain compression-based architectures may degrade clinically relevant information, particularly in advanced glaucoma.

The central challenge of visual field denoising lies in distinguishing measurement variability from true pathological scotomas. Compression-based models, including convolutional autoencoders and variational autoencoders, force data through a low-dimensional latent bottleneck and tend to reconstruct extreme values toward the population mean.^7^ In our study, these architectures consistently demonstrated reduced structure–function agreement relative to restoration-based methods. The CNN-VAE exhibited the greatest degradation, consistent with the possibility that probabilistic latent regularization attenuates localized deep defects.

When stratified by disease severity, structure–function concordance declined across all models in advanced glaucoma. This reduction was observed even in the raw baseline and is consistent with the well-described RNFLT floor effect,^15,16^ whereby structural thickness approaches a lower asymptote and additional functional loss is not linearly reflected in OCT measurements. Under these biological constraints, denoising did not improve prediction in severe disease. Notably, compression-based models demonstrated disproportionate degradation, supporting concern that latent bottleneck architectures may blunt deep and spatially localized scotomas.

By contrast, restoration-based architectures trained with pathology-aware objectives demonstrated greater stability across severity strata. Pathology-aware NAFNet achieved the highest global structure–function concordance and maintained competitive performance across borderline, mild, and moderate subgroups. By explicitly weighting reconstruction errors at severely depressed functional locations and clinically informative spatial regions, this strategy mitigated the tendency of conventional denoisers to attenuate pathological extremes. Importantly, however, even this optimized approach yielded only modest improvements over the raw baseline.

A key methodological insight emerged from our evaluation-direction sensitivity analysis. When prediction was performed in the RNFLT-to-VF direction, spatially averaged visual fields achieved performance comparable to, and in some settings exceeding, spatially preserved inputs. This indicates that RNFLT-to-VF mapping is largely driven by global disease severity rather than spatial defect morphology, limiting its utility for assessing whether denoising preserves localized glaucomatous patterns. In contrast, VF-to-RNFLT mapping penalized spatial collapse: replacing each visual field with its mean sensitivity value consistently reduced structure–function concordance. This asymmetry likely reflects the anatomical organization of the retinal nerve fiber layer, which encodes spatially specific bundle trajectories.^8,9^ Accordingly, evaluation in the VF-to-RNFLT direction provides a more spatially sensitive and biologically constrained assessment of denoising performance.

The limited magnitude of improvement observed even with pathology-aware restoration likely reflects fundamental biological constraints. The structure–function relationship in glaucoma is inherently variable due to inter-individual differences in optic disc position, nerve fiber bundle trajectories, and retinal ganglion cell distribution. The observed R^2^ ceiling of approximately 0.55 suggests that a substantial proportion of prediction error arises from anatomical mismatch rather than perimetric noise alone. Consequently, denoising can improve predictive performance only within these biological constraints.

We present a biologically grounded evaluation framework for visual field denoising based on structure–function mapping. Our findings demonstrate that visually smoother fields are not inherently more accurate representations of glaucomatous damage. Restoration-based models, particularly those incorporating pathology-aware training objectives, can modestly reduce variability while preserving biologically relevant functional signals. In contrast, compression-based architectures risk attenuating clinically meaningful defects, especially in advanced disease. Importantly, evaluation direction matters. RNFLT-to-VF mapping may overestimate denoising performance because it is dominated by global severity signals and does not require preservation of spatial defect morphology. In contrast, VF-to-RNFLT mapping provides a spatially sensitive validation framework that penalizes loss of localized patterns.

These results underscore the importance of validating artificial intelligence–based denoising tools against structural biomarkers in anatomically appropriate directions rather than relying solely on visual appearance, severity regression, or test–retest repeatability. In advanced glaucoma, where structural measurements approach a floor effect, raw visual field data remains a robust representation of functional loss, and aggressive denoising should be applied with caution.

## Data Availability

The data analyzed in this study are not publicly available due to patient privacy, institutional review board requirements, and data use restrictions. De-identified data may be available from the corresponding author upon reasonable request, subject to institutional approval and execution of appropriate data use agreements.

## Abbreviations

VF: visual field
RNFLT: retinal nerve fiber layer thickness
RNFL: retinal nerve fiber layer
RGC: retinal ganglion cell
AI: artificial intelligence
OCT: optical coherence tomography
TD: total deviation
SAP: standard automated perimetry
MAE: mean absolute error
MSE: mean squared error
RMSE: root mean square error
MD: mean deviation
HFA: Humphrey Field Analyzer
MEEI: Massachusetts Eye and Ear Infirmary
NIH: National Institutes of Health
CNN: convolutional neural network
AE: autoencoder
VAE: variational autoencoder
CNN-AE: CNN autoencoder
CNN-VAE: CNN variational autoencoder
NAFNet: Nonlinear Activation Free Network
N2N: Noise2Noise
N2V: Noise2Void
PosEnc: Positional Encoding Autoencoder

