## Supplemental Figure 1 for "Developing and Evaluating Deep Learning Approaches for Visual Field Denoising in Glaucoma"

**Supplemental Figure 1**. Formal mathematical explanation of the pathology-aware weighted reconstruction

Formally, the pathology-aware weighted reconstruction loss was defined as:

$$\mathcal{L}_{\text{weighted}} = \frac{1}{N}\sum_{i=1}^{N}\left(w_i^{\text{spatial}} \cdot w_i^{\text{severity}}\right)(y_i - \hat{y}_i)^2,$$

where $y_i$ and $\hat{y}_i$ denote the ground-truth and reconstructed total deviation (TD) values at test location $i$, and $N$ is the number of valid spatial locations.

The spatial prior $w_i^{\text{spatial}}$ is a fixed anatomical weighting matrix emphasizing clinically informative regions (e.g., central and arcuate zones). The severity-dependent term was defined as:

$$w_i^{\text{severity}} = \max(\sigma(-y_i), 0.1),$$

where $\sigma(\cdot)$ denotes the sigmoid function. This formulation increases loss contribution from severely depressed locations (large negative TD values) while enforcing a minimum weight to prevent underweighting of relatively preserved regions.
